# Frequency of and factors associated with antiseizure medication discontinuation discussions and decisions in patients with epilepsy: a multicenter retrospective chart review

**DOI:** 10.1101/2022.12.28.22283991

**Authors:** Samuel W Terman, Geertruida Slinger, Adriana Koek, Jeremy Skvarce, Mellanie V Springer, Julie M Ziobro, James F Burke, Willem M Otte, Roland D Thijs, Kees PJ Braun

## Abstract

**Objective:** Guidelines suggest considering antiseizure medication (ASM) discontinuation in patients with epilepsy who become seizure-free. Little is known about how discontinuation decisions are being made in practice. We measured the frequency of, and factors associated with, discussions and decisions surrounding ASM discontinuation.

**Methods:** We performed a multicenter retrospective cohort study at the University of Michigan (UM) and two Dutch centers: Wilhelmina Children’s Hospital (WCH) and Stichting Epilepsie Instellingen Nederland (SEIN). We screened all children and adults with outpatient epilepsy visits in January 2015 and included those with at least one visit during the subsequent two years where they were seizure-free for at least one year. We recorded whether charts documented 1) a discussion with the patient about possible ASM discontinuation and 2) any planned attempt to discontinue at least one ASM. We conducted multilevel logistic regressions to determine factors associated with each outcome.

**Results:** We included 1,058 visits from 463 patients. Of all patients who were seizure-free at least one year, 248/463 (53%) had documentation of any discussion and 98/463 (21%) planned to discontinue at least one ASM. Corresponding frequencies for patients who were seizure-free at least two years were 184/285 (65%) and 74/285 (26%). The probability of discussing or discontinuing increased with longer duration of seizure-freedom. Still, even for patients who were ten years seizure-free, our models predicated that in only 49% of visits was a discontinuation discussion documented, and in only 16% of visits was it decided to discontinue all ASMs. Provider-to-provider variation explained 18% of variation in whether patients discontinued any ASM.

**Significance:** Only approximately half of patients with prolonged seizure-freedom had a documented discussion about ASM discontinuation. Discontinuation was fairly rare even among low-risk patients. Future work should further explore barriers to and facilitators of counseling and discontinuation attempts.

**Key points box:**

- We performed a multicenter cohort study evaluating factors associated with discussions and decisions to discontinue antiseizure medications (ASMs).
- Of all patients seizure-free at least one year, 53% had documentation of any discussion and 21% planned to discontinue at least one ASM. Corresponding frequencies for patients seizure-free at least two years were 65% and 26%.
- While discussions and discontinuations increased with increasing seizure-free interval, even for patients who were ten years seizure-free, in only 49% of visits did providers discuss the possibility of discontinuation, and in only 16% of visits did patients decide to discontinue all ASMs.
- Provider-to-provider variation explained 18% of variation in whether patients discontinued any ASM.
- ASM discontinuation was fairly rare even among low-risk patients. Future work should further explore barriers to and facilitators of counseling and discontinuation attempts, including the role that differences in physician counseling play in determining whether patients discontinue.

## Introduction

Over 50 million people have epilepsy.^1^ Fortunately, antiseizure medications (ASMs) render two-thirds of patients seizure-free.^2^ For this group, a central question is whether ASMs are necessary indefinitely. ASMs reduce morbidity and mortality by reducing seizures.^3,4^ However, ASMs exert side effects^5,6^ which reduce quality of life,^7–12^ and relapse risk declines with increasing seizure-freedom.^13,14^ Accordingly, guidelines have endorsed considering ASM discontinuation after attaining seizure-freedom after detailed counseling.^15–17^

Despite literature estimating post-withdrawal relapse risk,^14^ little is known about real-world withdrawal decisions. One single-center study found that only 32% of seizure-free adult patients had discussed ASM withdrawal in the past year.^18^ Thus, we hypothesize that counseling regarding the possibility of withdrawal is rarer than suggested by past guidelines. However, that study included only self-reported patient data, was not stratified by key variables (e.g., seizure-free duration, post-discontinuation risk), and did not include children. Literature exploring the determinants of deciding to withdraw ASMs is sparse.^19,20^ One survey documented variation in how likely clinicians were to endorse discontinuing ASMs in response to several patient vignettes.^21^ Yet, hypothetical intent may not translate into practice, and selected vignettes reflect a small portion of real-world complexity. While there is no known ‘optimal’ time for patients to consider a discontinuation attempt, further investigation into the frequency of and factors associated with ASM discontinuation discussions and actual discontinuation plans would delineate whether clinicians are proceeding to withdrawal attempts in the lowest-risk patients.

We measured the frequency of, and factors associated with, documented discussions surrounding ASM discontinuation and plans to discontinue ASMs in patients with well-controlled epilepsy. We evaluated how discussions and decisions changed across clinically relevant covariates.

## Methods

### Study design and dataset

This was a retrospective cohort study. We abstracted information from electronic medical records of patients seen at 1) Stichting Epilepsie Instellingen Nederland (SEIN), 2) the University of Michigan (UM), and 3) University Medical Center Utrecht, Wilhelmina Children’s Hospital (WCH).

### Procedures involving human subjects

This study was deemed exempt from review by the University of Michigan and University Medical Center Utrecht Institutional Review Boards. Consent was not required.

### Patient selection

Patients were screened if they had at least one outpatient visit for epilepsy (International Classification of Disease-9 codes 345.xx), in January 2015. We began our observation period in 2015 to understand withdrawal decisions in practice without the use of the now-available individualized post-discontinuation seizure-risk calculator (introduced mid-2017).^22,23^ Also, as there was no way to know from these data which clinicians might be using the calculator on which patients, thus no way to adjust for or stratify by that variable, we wished to eliminate that unknowable/uncapturable source of heterogeneity.

We reviewed charts to confirm a diagnosis of epilepsy, according to International League Against Epilepsy definitions.^24^ We further restricted the sample to patients 1) at least 1 year of age (given that neonatal seizures represent a distinct clinical entity) with no age maximum, 2) with at least one visit where they were at least one year seizure-free in the subsequent two years of follow-up (1/2015 to 1/2017), 3) without previous epilepsy surgery, infantile spasms, juvenile myoclonic epilepsy, or childhood epileptic encephalopathies (scenarios with poorer prognosis, less relevant to ASM discontinuation), and 4) taking at least one ASM. Note that WCH included only children. While guidelines published before this study period suggested considering discontinuation after two-years seizure-free,^15,16^ we included patients at least one-year seizure-free 1) to better ensure we were not missing discussions before the two-year mark to prepare for future discontinuation decisions, 2) so we could more widely explore the effect of seizure-free durations given the possibility that some patients might be discontinuing before guideline-recommended timepoints, and 3) because risk decreases gradually with increasing seizure-free durations rather than abruptly at two years.

### Variables

We had two co-primary outcomes, which we adjudicated based on reviewing the medical decision-making portion of each clinic note within our study period. The first co-primary outcome was whether the electronic medical record documented a discussion with the patient surrounding the possibility of ASM discontinuation. For example, if a note stated “continue ASMs” without further explanation regarding reasons for or against and did not describe that a risk-benefit discussion occurred with the patient, such notes were counted as not having documented a discussion with the patient. The second co-primary outcome was whether the chart documented the plan to attempt any ASM discontinuation without intent to cross-taper onto a different agent. To determine these outcomes, we reviewed all office visits within two years after the first eligible visit (1/2015-1/2017).

We collected factors that may influence decision-making. These included basic demographic information (e.g., age, sex, race) and seizure risk predictors (Table 2).^22^ To test how each outcome varied according to overall risk, which contains more information at once than any single variable alone, we calculated each patient’s two-year post-withdrawal seizure risk.^22,23^ We also studied variables not captured in the calculator including seizure semiology (impairing awareness; motor), etiology, prior discontinuation attempts (as determined by reviewing visits before the first eligible visit), and provider characteristics at each visit (M.D./D.O. versus Nurse Practitioner/Physician Assistant; epilepsy specialist). We did not specifically collect discontinuation discussions prior to 2015, as our focus was on capturing current discussions, and it would seem important to document discussions surrounding medical decision-making regarding whether to continue ASMs at each visit regardless of whether a prior discussion occurred. We did, however, collect information about past discontinuation attempts, because these imply that a patient who previously discontinued had a relapse that led to restart of ASM, which may influence the chance of considering a current discontinuation attempt.

We recorded the date of first seizure relapse between the patient’s first included visit (earliest 1/2015) and 1/2022, up to seven years of follow-up.

### Statistical analysis

We described univariate statistics for categorical data using frequencies, and continuous data using medians and interquartile ranges (IQR). Based on the first visit in the study window, we compared baseline characteristics according to whether patients discussed ASM discontinuation or planned to discontinue any/all ASMs at any visit using t-tests and Chi-squared tests.

To evaluate the association between each factor and the outcomes, we conducted mixed-effects logistic regressions, with a random intercept for each patient. We did so per-visit because while many variables remain constant between visits (e.g., etiology, focality), other variables could change between visits (e.g., age, seizure-free duration, new versus return visit, type of provider, number of ASMs). Additionally, outcomes could change between visits (e.g., a patient could decide to not discontinue at one visit but then do so at the next visit). Thus, we included one observation for every visit during the two years of follow-up where the patient was at least one year seizure-free and taking at least one ASM at the start of the visit. We performed one model where the outcome was whether an ASM discontinuation discussion occurred, and additional models where the outcome was whether the patient and physician decided to withdraw any or all ASMs. We adjusted for all covariates listed in Table 2. Continuous variables were entered as cubic polynomials. We specified a priori variables to highlight graphically due to their clinical importance – age at the visit, years of seizure-freedom, latest EEG with interictal epileptiform discharges, a previous discontinuation attempt, taking at least one older-generation ASM, epileptologist provider, and site. We computed adjusted and unadjusted predicted probabilities from our logistic regressions and displayed results using bar graphs with 95% confidence intervals (CIs). We did a separate model including calculated multivariable risk^22,23^ as the sole predictor. We did so 1) to avoid multicollinearity, and 2) because it is well-known that multivariable risk prediction is better able to capture heterogeneity between patients, more powerful, and less likely to capture false positives compared with any single variable at a time.^25^

To evaluate the amount of variation in each outcome due to provider differences, we repeated the above adjusted mixed effects logistic regressions, using a random intercept for each provider (rather than each patient), to compute the intraclass correlation coefficient (ICC).^26^ The ICC represents the percent of total variation in the outcome due to differences between providers, after adjusting for patient characteristics listed in Table 2.

We performed a qualitative analysis in which we abstracted text from each note explaining decisions. We classified text into categories as determined by consensus of two independent raters who resolved all disagreements by discussion and displayed this information as frequencies.

Finally, to help interpret the rate of ASM discontinuation, we calculated adjusted and unadjusted seizure survival curves for periods of continuation versus tapering. We did so via discrete time logistic regressions (Methods Supplement). We entered ASM discontinuation as a time-varying covariate that started out as “no” for all patients, and then updated to “yes” at the time of deciding to discontinue ASMs if applicable.

Data were analyzed using SAS version 9.4 (Cary, NC) and Stata version 16.1 (College Station, TX).

### Data availability statement

Statistical code for this project may be obtained from the authors. Data can only be shared with a data use agreement.

## Results

### Population description per patient

We included 1,058 visits from 463 patients who were seizure-free at least one year, of whom 285 were seizure-free at least two years (Table 1). We included 122 children (age under 18 years at their first visit; total 122/463 = 26%; SEIN: 56/342 = 16%; UM: 34/89 = 38%; WCH: 32/32 = 100%). The median age at the first included visit was 32 years (IQR 17-53), 48% were female, and the largest contributing center was SEIN (342/463 = 74%). Table 2 provides additional information describing our population.

**Table 1:** Flowchart and outcome frequencies, stratified by center. Note that “Discontinue all ASMs” is a subset of “Discontinue any ASM.” Likewise, “Discontinue any ASM” is a subset of “Discussion.” The unit of analysis in this Table is per patient, collapsed over all visits.

|  | SEIN | UM | WCH | Total |
| --- | --- | --- | --- | --- |
| **Flowchart** |  |  |  |  |
| Epilepsy visit 1/2015 | 1812 | 261 | 252 | 2325 |
| >1y seizure-free | 439 | 101 | 76 | 616 |
| No surgery | 396 | 97 | 51 | 544 |
| No other exclusions | 361 | 90 | 38 | 489 |
| At least 1 ASM | 342 | 89 | 32 | 463 |
| **Among all included patients, at least one year seizure-free at any visit** |  |  |  |  |
| Discussion | 192/342 (56%) | 35/89 (39%) | 21/32 (66%) | 248/463 (54%) |
| Discontinue any ASM | 69/342 (20%) | 15/89 (17%) | 14/32 (44%) | 98/463 (21%) |
| Discontinue all ASMs | 41/342 (12%) | 11/89 (12%) | 14/32 (44%) | 66/463 (14%) |
| Reduce dose but not discontinue | 19/342 (6%) | 2/89 (2%) | 1/32 (3%) | 22/463 (5%) |
| **Among all included patients, at least two years seizure-free at any visit** |  |  |  |  |
| Discussion | 144/210 (69%) | 28/59 (47%) | 12/16 (75%) | 184/285 (65%) |
| Discontinue any ASM | 53/210 (25%) | 12/59 (20%) | 9/16 (56%) | 74/285 (26%) |
| Discontinue all ASMs | 33/210 (16%) | 8/59 (14%) | 9/16 (56%) | 50/285 (18%) |
| Reduce dose but not discontinue | 1/210 (8%) | 1/59 (2%) | 1/16 (6%) | 19/285 (7%) |

ASM: antiseizure medication; SEIN: Stichting Epilepsie Instellingen Nederland; UM: University of Michigan; WCH: Wilhelmina Children’s Hospital

**Table 2:**
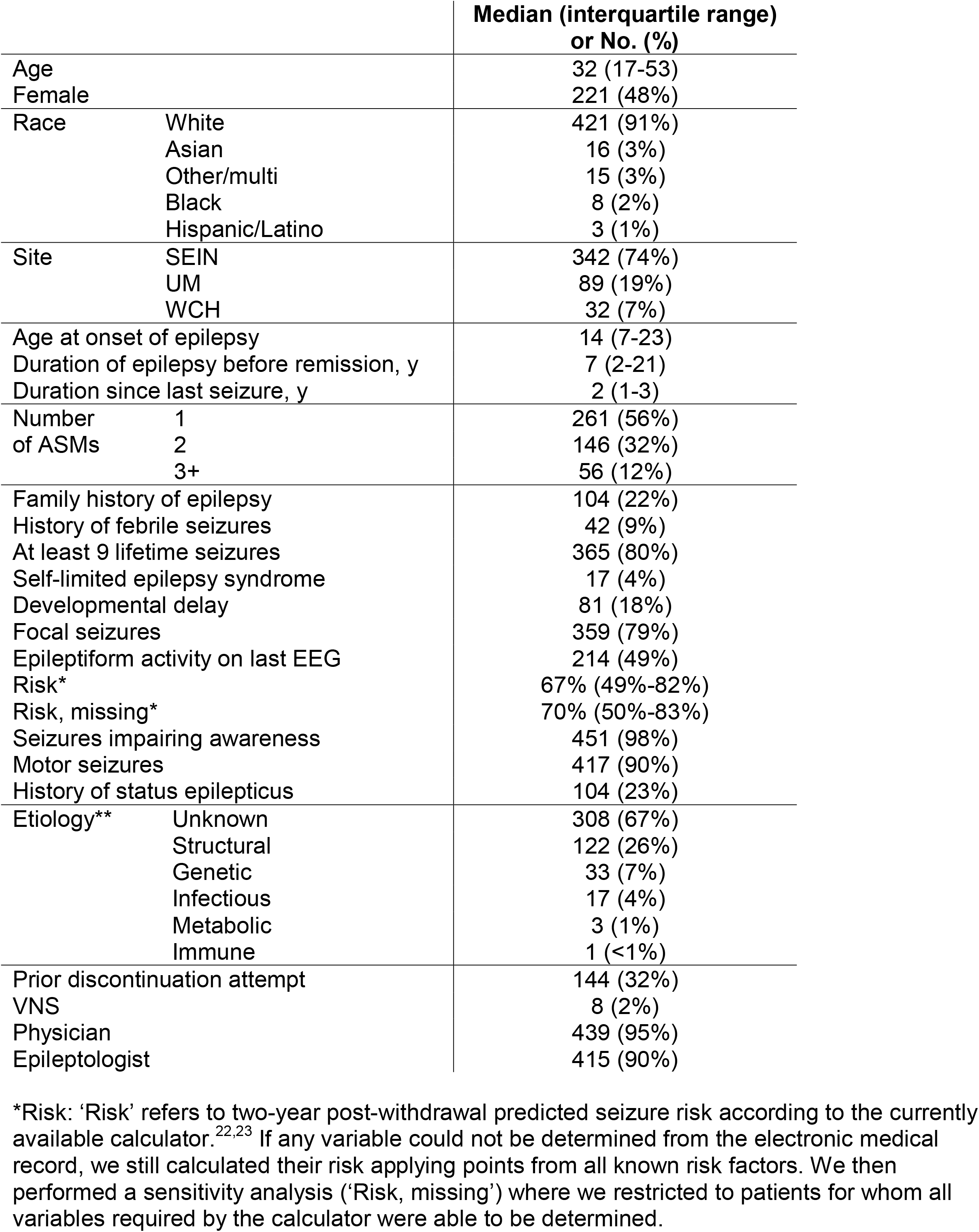
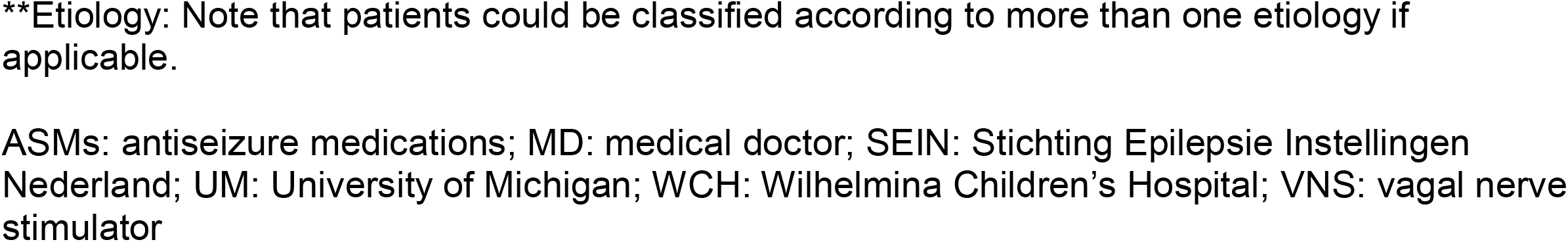
Baseline characteristics of all included patients. This is based on the first eligible visit for each patient, one datapoint per patient. N=463.

|  |  | Median (interquartile range)<br>or No. (%) |
| --- | --- | --- |
| Age |  | 32 (17-53) |
| Female |  | 221 (48%) |
| Race | White | 421 (91%) |
|  | Asian | 16 (3%) |
|  | Other/multi | 15 (3%) |
|  | Black | 8 (2%) |
|  | Hispanic/Latino | 3 (1%) |
| Site | SEIN | 342 (74%) |
|  | UM | 89 (19%) |
|  | WCH | 32 (7%) |
| Age at onset of epilepsy |  | 14 (7-23) |
| Duration of epilepsy before remission, y |  | 7 (2-21) |
| Duration since last seizure, y |  | 2 (1-3) |
| Number of ASMs | 1 | 261 (56%) |
|  | 2 | 146 (32%) |
|  | 3+ | 56 (12%) |
| Family history of epilepsy |  | 104 (22%) |
| History of febrile seizures |  | 42 (9%) |
| At least 9 lifetime seizures |  | 365 (80%) |
| Self-limited epilepsy syndrome |  | 17 (4%) |
| Developmental delay |  | 81 (18%) |
| Focal seizures |  | 359 (79%) |
| Epileptiform activity on last EEG |  | 214 (49%) |
| Risk* |  | 67% (49%-82%) |
| Risk, missing* |  | 70% (50%-83%) |
| Seizures impairing awareness |  | 451 (98%) |
| Motor seizures |  | 417 (90%) |
| History of status epilepticus |  | 104 (23%) |
| Etiology** | Unknown | 308 (67%) |
|  | Structural | 122 (26%) |
|  | Genetic | 33 (7%) |
|  | Infectious | 17 (4%) |
|  | Metabolic | 3 (1%) |
|  | Immune | 1 (<1%) |
| Prior discontinuation attempt |  | 144 (32%) |
| VNS |  | 8 (2%) |
| Physician |  | 439 (95%) |
| Epileptologist |  | 415 (90%) |

\*Risk: 'Risk' refers to two-year post-withdrawal predicted seizure risk according to the currently available calculator.<sup>22,23</sup> If any variable could not be determined from the electronic medical record, we still calculated their risk applying points from all known risk factors. We then performed a sensitivity analysis ('Risk, missing') where we restricted to patients for whom all variables required by the calculator were able to be determined.
**\*\*Etiology:** Note that patients could be classified according to more than one etiology if applicable.
ASMs: antiseizure medications; MD: medical doctor; SEIN: Stichting Epilepsie Instellingen Nederland; UM: University of Michigan; WCH: Wilhelmina Children's Hospital; VNS: vagal nerve stimulator

### Frequency of ASM discontinuation discussions and planned attempts per patient

Among patients who were seizure-free at least one year, 248 (54%) had a documented discussion at any visit regarding the possibility of discontinuation, 98 (21%) planned to discontinue any ASM, and of those 98, 66 (14% of the total sample) planned to discontinue all ASMs at any point during the two years of follow-up. Among patients who were seizure-free at least two years, 184 (65%) had any documented discussion regarding the possibility of discontinuation, 74 (26%) planned to discontinue any ASM, and of those 75, 50 (18% of the total sample) planned to discontinue all ASMs at any point during the two years of follow-up. Among 248 patients who had any documented discussion, 98 (40%) planned to discontinue any ASM (SEIN: 69/192 = 36%; UM: 15/35 = 43%; WCH: 14/21 = 67%).

Rates of discontinuing all ASMs, according to number of baseline ASMs, were: 1 baseline ASM: 53/261 (20%); 2 baseline ASMs: 10/146 (7%); 3 baseline ASMs: 3/56 (5%) (unadjusted p-value: <0.01; adjusted p-value: 0.03).

We followed the 66 patients who decided to discontinue all ASMs within the two-year observation period, until January 1, 2022, to determine how frequently patients actually discontinued all ASMs as planned. Charts documented that 45/66 (68%) completed discontinuation of all ASMs. Of the remaining 21, 5 had a seizure during tapering and thus resumed, 8 started tapering then subsequently decided against it despite no seizure, and 8 had inadequate follow-up to determine whether they completed discontinuation.

Among those 398 patients who did not discontinue all ASMs during the 2015-2017 observation period, 33 patients (33/398 = 8%) decided to discontinue all ASMs subsequently at some point before January 1, 2022.

Figure 1 displays what percent of patients decided to discontinue each ASM. For the most common ASMs (levetiracetam, valproate, carbamazepine, lamotrigine), this percent was between 10%-19%.

**Figure 1:**
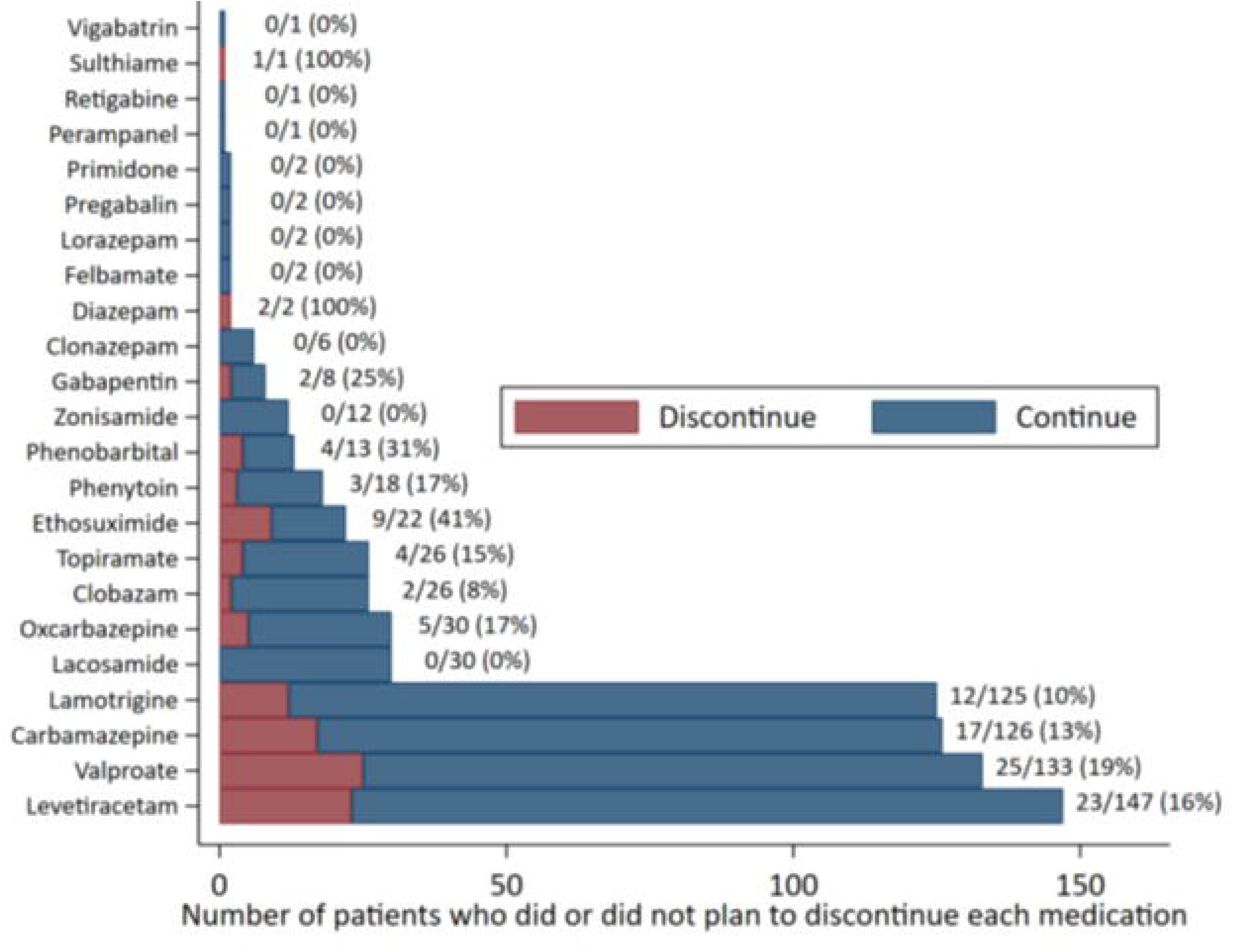
Frequency of planning to discontinue each antiseizure medication. There is one datapoint for each person-medication taken at any point during 2015-2017.

### Factors associated with ASM discontinuation discussions and attempts per visit

Variables that predicted an increased unadjusted chance of having a discontinuation discussion were: younger age, WCH site, longer duration of seizure-freedom, self-limited syndrome, absence of developmental delay, absence of interictal epileptiform EEG findings, and seeing a physician as opposed to a physician extender (all p<0.05; Table 3; Supplemental Figure 1). Significant adjusted predictors were: longer duration of seizure-freedom, older generation ASM, absence of developmental delay, and lower calculated risk (all p<0.05) (Figure 2).

**Table 3:** Baseline characteristics according to whether there was a documented discussion regarding discontinuation of any antiseizure medication (ASM) and whether patients planned to discontinue any ASM, at any visit during the two years of follow-up. Counts and percentages are per patient, collapsed across all visits. Unadjusted (Unadj.) p-values represent Chi-squared or t-tests per patient. Adjusted (Adj.) p-values represent multivariable mixed-effects logistic regressions per visit adjusted for all terms listed in Table 2, given we felt that it was also important to evaluate outcomes per-visit because both certain predictors and outcomes can change between visits. Continuous covariates are not presented here because they are instead presented in the Figures.

|  |  | Discussion |  |  | Discontinue any ASM |  |  |
| --- | --- | --- | --- | --- | --- | --- | --- |
|  |  | p-value |  |  | p-value |  |  |
|  |  | Number (%) | Unadj. | Adj. | Number (%) | Unadj. | Adj. |
| Demographics |  |  |  |  |  |  |  |
| Age |  | Figure 2 and<br>Supp. Figure 1 | **<0.01** | 0.72 | Figure 2 and<br>Supp. Figure 1 | **<0.01** | 0.25 |
| Sex | Male | 121/242 (50%) | 0.11 | 0.09 | 50/242 (21%) | 0.78 | 0.95 |
|  | Female | 127/221 (57%) |  |  | 48/221 (22%) |  |  |
| Race | White | 227/421 (54%) | 0.29 | 0.48 | 86/421 (20%) | 0.17 | 0.46 |
|  | Asian | 11/16 (69%) |  |  | 6/16 (38%) |  |  |
|  | Other/multi | 5/15 (33%) |  |  | 5/15 (33%) |  |  |
|  | Black | 3/8 (38%) |  |  | 0/8 (0%) |  |  |
|  | Hispanic/Latino | 2/3 (67%) |  |  | 1/3 (33%) |  |  |
| Site | SEIN | 192/342 (56%) | **<0.01** | 0.74 | 69/342 (20%) | **<0.01** | 0.58 |
|  | UM | 35/89 (39%) |  |  | 15/89 (17%) |  |  |
|  | WCH | 21/32 (66%) |  |  | 14/32 (44%) |  |  |
| Additional variables to calculate 2-year post-withdrawal seizure risk, from initial visit |  |  |  |  |  |  |  |
| Years since<br>last seizure |  | Figure 2 and<br>Supp. Figure 1 | **<0.01** | **<0.01** | Figure 2 and<br>Supp. Figure 1 | **<0.01** | **0.01** |
| Number<br>of baseline<br>ASMs* | 1 | 130/261 (50%) | 0.17 | 0.72 | 53/261 (20%) | 0.75 | 0.40 |
|  | 2 | 84/146 (58%) |  |  | 34/146 (23%) |  |  |
|  | 3+ | 34/56 (61%) |  |  | 11/56 (20%) |  |  |
| Older gen.<br>ASM | No | 98/198 (50%) | 0.13 | **0.01** | 35/198 (18%) | 0.11 | **0.02** |
|  | Yes | 150/265 (57%) |  |  | 63/265 (24%) |  |  |
| Family history<br>of epilepsy | No | 194/359 (54%) | 0.15 | 0.34 | 76/359 (21%) | >0.99 | 0.81 |
|  | Yes | 54/104 (52%) |  |  | 22/104 (21%) |  |  |
| Febrile<br>seizures | No | 222/421 (53%) | 0.26 | 0.74 | 86/421 (20%) | 0.22 | 0.68 |
|  | Yes | 26/42 (62%) |  |  | 12/42 (29%) |  |  |
| At least 9<br>seizures | No | 49/90 (54%) | 0.90 | 0.10 | 23/90 (26%) | 0.27 | 0.14 |
|  | Yes | 196/365 (54%) |  |  | 74/365 (20%) |  |  |
| Self-limited<br>syndrome | No | 231/440 (53%) | **0.02** | 0.08 | 83/440 (19%) | **<0.01** | **0.04** |
|  | Yes | 14/17 (82%) |  |  | 13/17 (76%) |  |  |
| Develop.<br>delay | No | 214/381 (56%) | **0.02** | **<0.01** | 80/381 (21%) | 0.81 | 0.30 |
|  | Yes | 34/81 (42%) |  |  | 18/81 (12%) |  |  |
| Focal<br>seizures | No | 44/95 (46%) | 0.13 | 0.29 | 21/95 (22%) | 0.71 | 0.67 |
|  | Yes | 198/359 (55%) |  |  | 73/359 (20%) |  |  |
| Epileptiform<br>EEG | No | 132/221 (60%) | **0.02** | 0.09 | 51/221 (23%) | 0.45 | 0.16 |
|  | Yes | 103/214 (48%) |  |  | 43/214 (20%) |  |  |
| Risk |  | Figure 2 | N/A | **<0.01** | Figure 2 | N/A | **<0.01** |
| Additional epilepsy characteristics |  |  |  |  |  |  |  |
| Impairing awareness | No | 5/11 (45%) | 0.59 | 0.92 | 3/11 (27%) | 0.62 | 0.53 |
|  | Yes | 242/451 (54%) |  |  | 95/451 (21%) |  |  |
| Motor seizures | No | 26/45 (58%) | 0.56 | 0.74 | 11/45 (24%) | 0.58 | 0.91 |
|  | Yes | 222/417 (53%) |  |  | 87/417 (21%) |  |  |
| Status epilepticus | No | 185/346 (53%) | 0.63 | 0.56 | 75/346 (22%) | 0.59 | 0.81 |
|  | Yes | 53/104 (51%) |  |  | 20/104 (19%) |  |  |
| Etiology | Unknown | 164/308 (53%) | 0.85 | Ref | 71/308 (23%) | 0.16 | Ref |
|  | Structural | 65/122 (53%) | 0.94 | 0.72 | 18/122 (15%) | **0.04** | 0.66 |
|  | Genetic | 14/33 (42%) | 0.18 | 0.32 | 7/33 (21%) | >0.99 | 0.20 |
|  | Infectious | 11/17 (65%) | 0.35 | 0.52 | 2/17 (12%) | 0.33 | 0.07 |
|  | Metabolic | 1/3 (33%) | 0.48 | 0.81 | 1/3 (33%) | 0.61 | 0.15 |
|  | Immune | 0/1 (0%) | 0.28 | * | 0/1 (0%) | 0.60 | ** |
| Prior disc. attempt | No | 168/312 (54%) | 0.94 | 0.89 | 63/312 (20%) | 0.41 | 0.36 |
|  | Yes | 77/144 (53%) |  |  | 34/144 (34%) |  |  |
| Provider characteristics, all visit |  |  |  |  |  |  |  |
| Physician | No | 6/24 (25%) | **<0.01** | 0.21 | 2/24 (8%) | 0.11 | **0.03** |
|  | Yes | 242/439 (55%) |  |  | 96/439 (22%) |  |  |
| Epileptologist | No | 24/48 (50%) | 0.60 | 0.31 | 17/48 (35%) | **0.01** | 0.86 |
|  | Yes | 224/415 (54%) |  |  | 81/415 (20%) |  |  |

\*Results were as follows for whether patients discontinued all ASMs, according to number of baseline ASMs: 1 baseline ASM: 53/261 (20%); 2 baseline ASMs: 10/146 (7%); 3 baseline ASMs: 3/56 (5%); unadjusted p-value: <0.01; adjusted p-value: 0.03.
\*\*Omitted from the adjusted model given collinearity.
ASMs: antiseizure medications; MD: medical doctor; SEIN: Stichting Epilepsie Instellingen Nederland; UM: University of Michigan; WCH: Wilhelmina Children's Hospital

**Figure 2:**
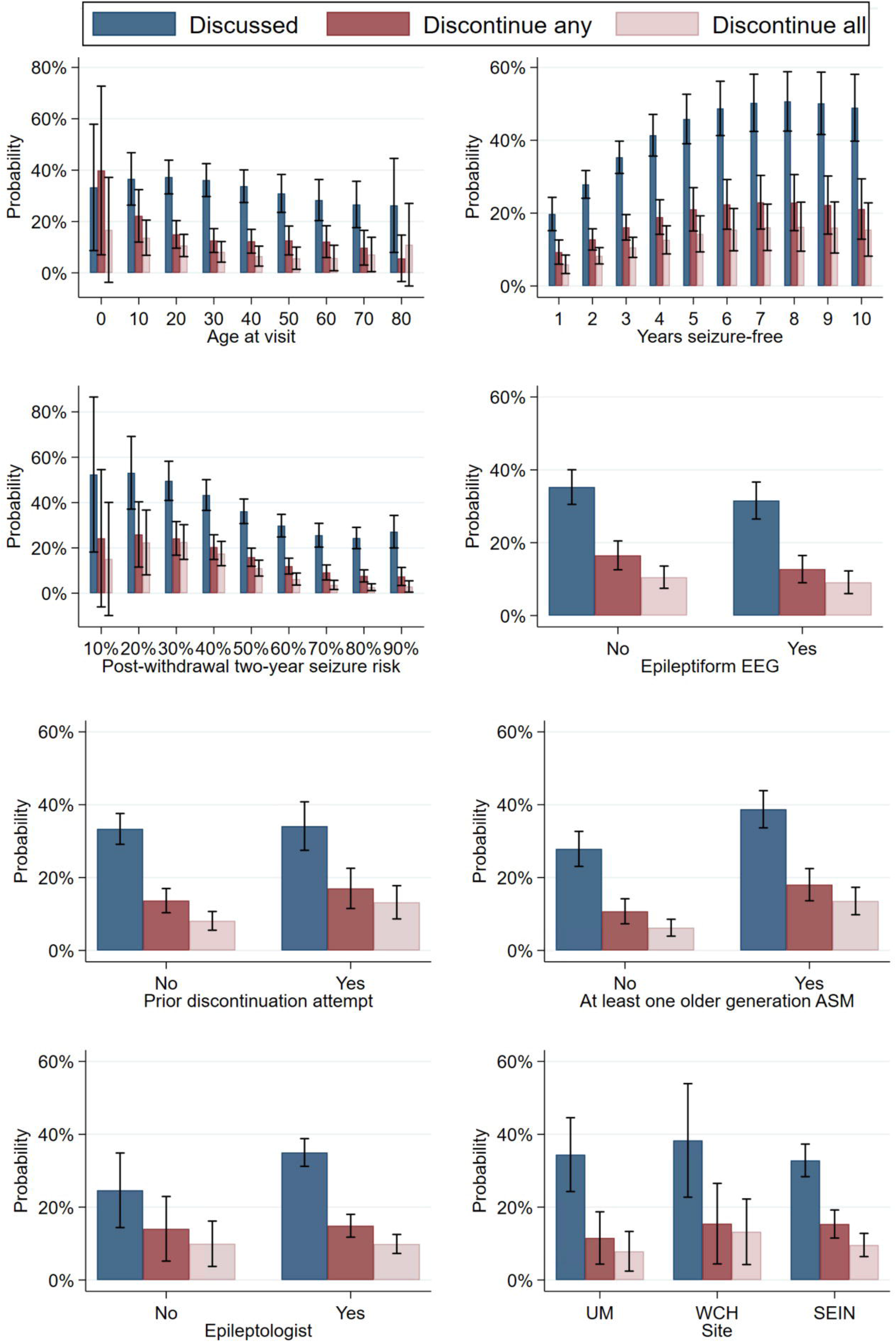
Adjusted probabilities and 95% confidence intervals of discussing or planning to discontinue any or all antiseizure medications (ASMs), stratified by a priori clinical variables. Each visit represented an observation.

Variables that predicted an increased unadjusted chance of planning to discontinue any ASM were: younger age, WCH site, longer duration seizure-freedom, self-limited syndrome, non-structural etiology, and seeing a non-epileptologist (all p<0.05). Significant adjusted predictors were: longer duration seizure-freedom, older generation ASM, self-limited syndrome, lower calculated risk, and seeing a physician (all p<0.05).

Given the importance of driving in clinical decisions, we stratified outcomes according to whether patients were above the legal driving age at their first eligible visit (Supplemental Table 1). Results similarly suggested that younger age predicted an unadjusted increased chance of discontinuation but was not significantly associated with discontinuation attempts after adjustment for all other variables and was not significantly associated with discussions.

As depicted in Figure 2, the probability of discussing discontinuation and attempting to discontinue increased with duration of seizure-freedom and decreased with increasing calculated post-withdrawal risk. Still, even for patients who were ten years seizure-free, in only 49% of visits did our models predict a discussion, and in only 16% of visits did our models predict that patients decided to discontinue all ASMs. Likewise, our models predicted that for patients with a two-year post-withdrawal relapse risk of 10% (i.e., a very low-risk patient), in only 52% of visits did providers document that they discussed the possibility of discontinuation and in only 15% of visits was it decided to discontinue all ASMs.

Provider-to-provider variation explained 11% (the ICC) of adjusted variation in whether a discussion was documented and 18% of whether any discontinuation was attempted (Supplemental Table 2).

### Reasons for ASM discontinuation decisions per patient

There were 257 patients for whom at least one note explained the decision whether to discontinue any ASM (Table 4). The most commonly stated reasons influencing decisions were seizure-free duration, fear of seizure relapse, and chance of another seizure. Seizure-free duration was cited as the most common reason to discontinue ASMs (i.e., due to decreasing relapse risk), but also as the most common reason to continue ASMs (i.e., due to satisfaction with seizure-freedom on current treatment).

**Table 4:** Qualitative reasons provided in electronic medical record clinic visit assessments underlying why patients did or did not plan to discontinue antiseizure medications (ASMs). For example, “side effects” refers to whether an explicit mention in the chart was made regarding the presence or absence of side effects as a motivating reason to discontinue or not. While this table contains 257 patients, the total N=450 is greater than 257 given each patient may have had more than one factor contributing to the decision whether to discontinue any ASM. The unit of analysis in this Table is per patient, collapsed over all visits.

| Factor | Discontinue any ASM |  |  |
| --- | --- | --- | --- |
|  | Total<br>N=450 | Yes<br>N=179 | No<br>N=271 |
| Seizure-free duration | 141 | 84 | 57 |
| Fear of seizure relapse | 45 | 9 | 36 |
| Chance of another seizure | 33 | 11 | 22 |
| Abnormal EEG | 30 | 19 | 11 |
| Psychosocial | 29 | 8 | 21 |
| Side effects | 29 | 8 | 21 |
| Previous unsuccessful wean | 22 | 1 | 21 |
| Driving | 20 | 1 | 19 |
| Other | 19 | 10 | 9 |
| School/work | 13 | 6 | 7 |
| ASM co-indication | 8 | 4 | 4 |
| Low-dose | 8 | 6 | 2 |
| Unclear whether events are seizures* | 6 | 1 | 5 |
| Psychiatric reasons | 6 | 2 | 4 |
| Pregnancy | 6 | 3 | 3 |
| Other medical condition | 5 | 1 | 4 |
| MRI abnormality | 5 | 1 | 4 |
| Previous status epilepticus | 4 | 1 | 3 |
| Upcoming surgery | 3 | 1 | 2 |

\*All patients in this cohort were determined seizure-free at least one year. However, patients in this group temporarily experienced clinically ambiguous events leading to clinical uncertainty their events were eventually evaluated to be non-epileptic.

### Seizure relapse cumulative incidence

Supplemental Figure 2 displays our population’s cumulative incidence of having at least one seizure relapse after the first eligible visit, according to whether at least one ASM, or all ASMs, were planned for discontinuation. For example, the adjusted cumulative incidence within one year of the initial visit was 15% (95% CI 11%-18%) during ‘continuation’ periods versus 38% (95% CI 20%-57%) for periods after deciding to discontinue all ASMs. Cumulative incidences by two years were 27% (95% CI 22%-31%) versus 53% (95% CI 37%-69%).

## Discussion

Among patients with epilepsy attaining at least one year of seizure-freedom, about half of patients had a documented discussion about ASM discontinuation, and 14% planned to discontinue all ASMs. Our data suggest that robust counseling regarding the pros and cons of discontinuation, or at least electronic medical record documentation explaining the patient’s tailored recommendations, may be less common than optimal.

There are many reasons clinicians and patients may be reluctant to withdraw ASMs, thus discontinuation may not be the best option for all patients. For a patient who feels that their ASMs have been helpful and well-tolerated, it may be quite sensible to continue long-term treatment. Furthermore, while a freely available rapid point-of-care post-withdrawal seizure risk calculator now exists, demonstrating moderate discrimination during development^22^ and external validation in several studies,^27,28^ another study found poor external validation,^29^ and clinicians still lack a unified robust model predicting both individualized continuation and discontinuation risks. Thus, while our data suggest ASM discontinuation is somewhat rare, certainly caution is required before pursuing discontinuation.

Nonetheless, our results highlight a potential asymmetry in epilepsy care. Epilepsy is diagnosed, and an ASM likely initiated, after a patient’s 10-year seizure risk exceeds 60%.^24^ However, in our dataset, even when the two-year post-withdrawal seizure relapse risk was predicted to be as low as 10% (which corresponds to a far less than 60% ten-year risk), in only 15% of visits did patients and their clinicians elect to attempt discontinuation. This result is only reinforced by external validation literature suggesting that the current risk calculator may overpredict risk,^27,28^ thus such patients could have been even lower risk than predicted. Existing literature does not inform below what seizure risk patients should consider discontinuation. Still, our results suggest the possibility that discontinuation decisions may be too conservative relative to standard of care when deciding whether to initiate ASMs.

Some previous work has examined factors associated with ASM withdrawal.^19,20^ For example, one study^20^ found an increased chance of treatment discontinuation in children, and cases with cryptogenic etiologies, fewer seizures, and normal neuroimaging. Our study builds upon those findings by adding in adjusted estimates, adding in calculated risk using multimodal predictors rather than only single dimensions like age or seizure-free duration, and simultaneously evaluating discontinuation discussions and decisions.

Younger age, self-limited syndromes, and treatment at WCH (children-only) predicted an unadjusted increased chance of withdrawal attempts. Children with self-limited epilepsy syndromes may have a favorable post-withdrawal prognosis appropriately prompting clinicians to consider eventual discontinuation, and withdrawal decisions have fewer implications for driving privileges in children. Age and site were not significant after adjusting for self-limited syndromes, suggesting that age itself may not be the only relevant factor after considering the remainder of a patient’s risk profile. Still, we found a gradual decline in withdrawal attempts with age, rather than an abrupt cutoff at ages 16 (US) to 18 (Netherlands) which might have been expected if decisions purely surrounded driving. It was also interesting that discontinuation was so rare in older patients, which echoes prior work.^19^ ASMs have special implications for aging-related comorbidities given ASMs are associated with a relative risk of falls,^30^ enzyme-inducing ASMs increase osteoporosis^31^ and lipids,^32,33^ and ASMs may worsen cognition throughout life^5,34^ beyond the known bidirectional relationship between epilepsy itself and dementia.^35^ Still, caution may be appropriate in older patients given unique psychosocial concerns regarding injury or fall risk from seizures in the context of declining bone health or potential anticoagulation or fear of losing independence, etc. Thus, our work provides background for future studies investigating the degree to which this represents appropriate caution versus frequent overtreatment with increasing age.

It was also interesting that EEG abnormalities, previous status epilepticus, or previous discontinuation attempts did not predict discontinuation decisions nearly as much as we had hypothesized. For example, it is possible that most patients or clinicians have already decided their course of action before ordering the EEG, or else a patient’s life circumstances, or other preferences may underly counseling to a greater extent than would EEG findings. These data might argue against frequent ordering of EEGs to inform withdrawal decisions if they ultimately have little influence on decision-making beyond all other factors that we captured.

Provider-to-provider variation accounted for 11%-18% of whether there was a discontinuation discussion or planned attempt. Physicians were twice as likely as non-physician practitioners to document discussion of possible discontinuation, and nearly three times more likely to pursue discontinuation. How a provider presents risks and benefits to a patient may critically influence patient decisions. Limited work in ASM withdrawal has documented the possibility of ‘framing effects’ whereby a patient might be more inclined to withdraw if presented with information in terms of ‘seizure-free probability’ rather than ‘seizure probability.’^36^ Future work should seek to better understand how patients wish to receive information regarding seizure risk and treatment benefits and harms, how risk communication practices vary between providers, and how to identify optimal risk communication techniques.

Finally, it was interesting that when a reason was provided by the electronic medical record explaining decisions, seizure-free duration was the most cited factor favoring both continuation and discontinuation (Table 4). Increasing seizure-free duration is likely a marker for a lower-risk patient, thus could favor discontinuation. However, seizure-free duration could also be an argument for continuation (e.g., given seizure-freedom and perceived therapeutic benefit this long on ASMs, “don’t rock the boat”). This highlights a tension: the same patient characteristic may be an argument for either decision, depending on how it is interpreted within a patient’s unique clinical context. Previous ASM discontinuation guidelines recommended considering discontinuation after a minimum of two years of seizure-freedom.^15,16^ However, if longer durations of seizure-freedom actually represent a factor in favor of continuation for some, guidelines based predominantly on time-based cutoffs may be overly simplistic and not capture important clinical nuances. Ideally, future guidelines would be based upon what ranges of risk, computed by multivariable techniques, predict improved quality of life, within the framework of a patient’s individual preferences and risk tolerance.

Our work has limitations. Charts may not capture the full extent of patient-physician conversations or counseling, charts do not inform whether discussions were patient-versus provider-initiated, and it is not possible to capture every single factor potentially relevant to decision-making. However, charts represent the gold standard data source short of recording and transcribing clinic encounters to understand the rationale for decisions, and while charts could undercount discussions it seems likely that charts accurately document all planned discontinuation attempts made at each visit. Future prospective data collection could address such limitations. We captured discontinuation attempts prior to our study window but may have missed previous discussions. Regardless, it is important to document the full extent of medical decision making and counseling at each visit. Our sample also consisted of neurology providers in specialized centers, several of which have been on the forefront of ASM discontinuation research. Thus, discussions or discontinuation attempts could be less common in less specialized settings less familiar with the possibility or process of discontinuation. In contrast, patients at our centers may be complex with longer durations more a larger number of ASM attempts until remission, thus it is possible that discontinuation attempts may be more common in community-based samples.” We also did not assess the influence of introducing the currently available post-withdrawal risk calculator^22,23^ on discussions or decisions. Comparing outcomes “pre versus post” calculator introduction within a single cohort would have been difficult to interpret, as each patient would inherently be in a different time-point in their disease course, and uptake of the calculator by clinicians was unknown from charts. Next, our survival curves likely overestimated seizure relapse rates given the lowest-risk patients may have the shortest follow-up time in a specialized center. Still, we reproduced a similar relative effect size compared with existing randomized data.^37,38^ Survival curves were also not individualized. Our goal in this manuscript was to model factors influencing discontinuation discussions and decisions, and our future work will further refine individualized seizure prediction. Next, no study can capture all possible sources of heterogeneity driving decisions, and higher-order interactions may exist in the real world. Nevertheless, we believe that we have captured the major sources of heterogeneity (e.g., age, etiology, syndrome, etc.), we had no clear a priori reason to suspect any specific interaction, and with an already complex model caution is needed when interpreting multiple comparisons. We included multivariable risk as a predictor to address this limitation – while we used the most comprehensive and rigorous available calculator,^22,23^ we recognize that risk estimates are imperfect.^27–29^

## Conclusions

Only approximately half of seizure-free patients had a documented discussion about possible future ASM discontinuation, and even in low-risk patients discontinuation was relatively rare. Differences between providers explained a substantial amount of variation in whether patients decided to discontinue ASMs, even after adjusting for many patient factors, which should prompt future work exploring differences in provider communication methods. Guidelines should not base discontinuation recommendations solely on seizure-free duration, as increasing seizure-free duration may be an argument to either discontinue (decreasing risk) or continue (marker of increased perceived efficacy) depending on the patient. Rather, guidelines should take a more nuanced multivariable approach. Future research is needed to understand barriers and facilitators to discontinuation and identify optimal risk thresholds to consider ASM discontinuation.

## Ethical publication statement

We confirm that we have read the Journal’s position on issues involved in ethical publication and affirm that this report is consistent with those guidelines.

## Author contributions

S.W.T. conceived of and designed the study, collected data, executed the statistical analysis, and wrote the manuscript. G.S. designed the study and collected and interpreted the data. A.K., J.S. and M.V.S collected data. All authors edited the manuscript. J.F.B., R.D.T., and K.PJ.B. provided study supervision.

---

**Supplemental Table 1:**
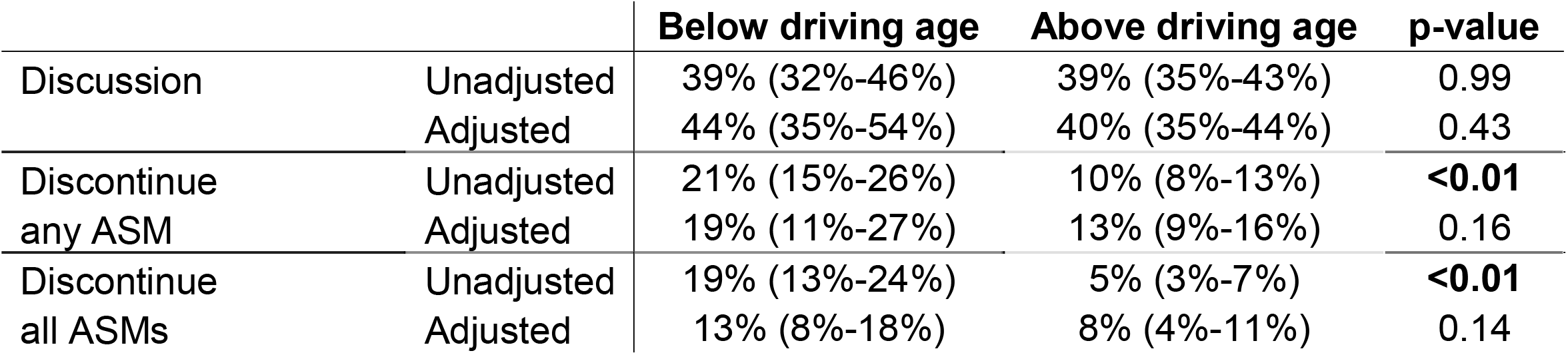
Probability of discontinuation discussions and planning to discontinue at least one antiseizure medication (ASM) according to being below versus above the legal driving age at the time of the first visit in our observation period (≥16 years in the US; ≥18 years in the Netherlands). Each visit represented an observation.

**Supplemental Table 2:** Percent variation in each outcome due to provider-to-provider differences. Each visit represented an observation. Each number displays the intraclass correlation coefficient (ICC) and 95% confidence interval from multilevel logistic regressions where the outcome was either discussion (top row) or discontinuation (bottom row), with a random intercept for each provider. We adjusted for all variables listed in Table 2. Provider characteristics were whether the provider is an MD versus physician extender, and epileptologist versus non-epileptologist.

|  | **ICC,**<br>**unadjusted** | **ICC, adjusted for**<br>**patient characteristics** | **ICC, adjusted for patient +**<br>**provider characteristics** |
| --- | --- | --- | --- |
| **Discussion** | 12% (6%-22%) | 11% (5%-23%) | 11% (5%-22%) |
| **Discontinue any ASM** | 20% (10%-37%) | 19% (9%-37%) | 18% (8%-35%) |
| **Discontinue all ASMs** | 26% (12%-46%) | 14% (4%-38%) | 13% (8%-38%) |

ASM: antiseizure medication

**Supplemental Figure 1:**
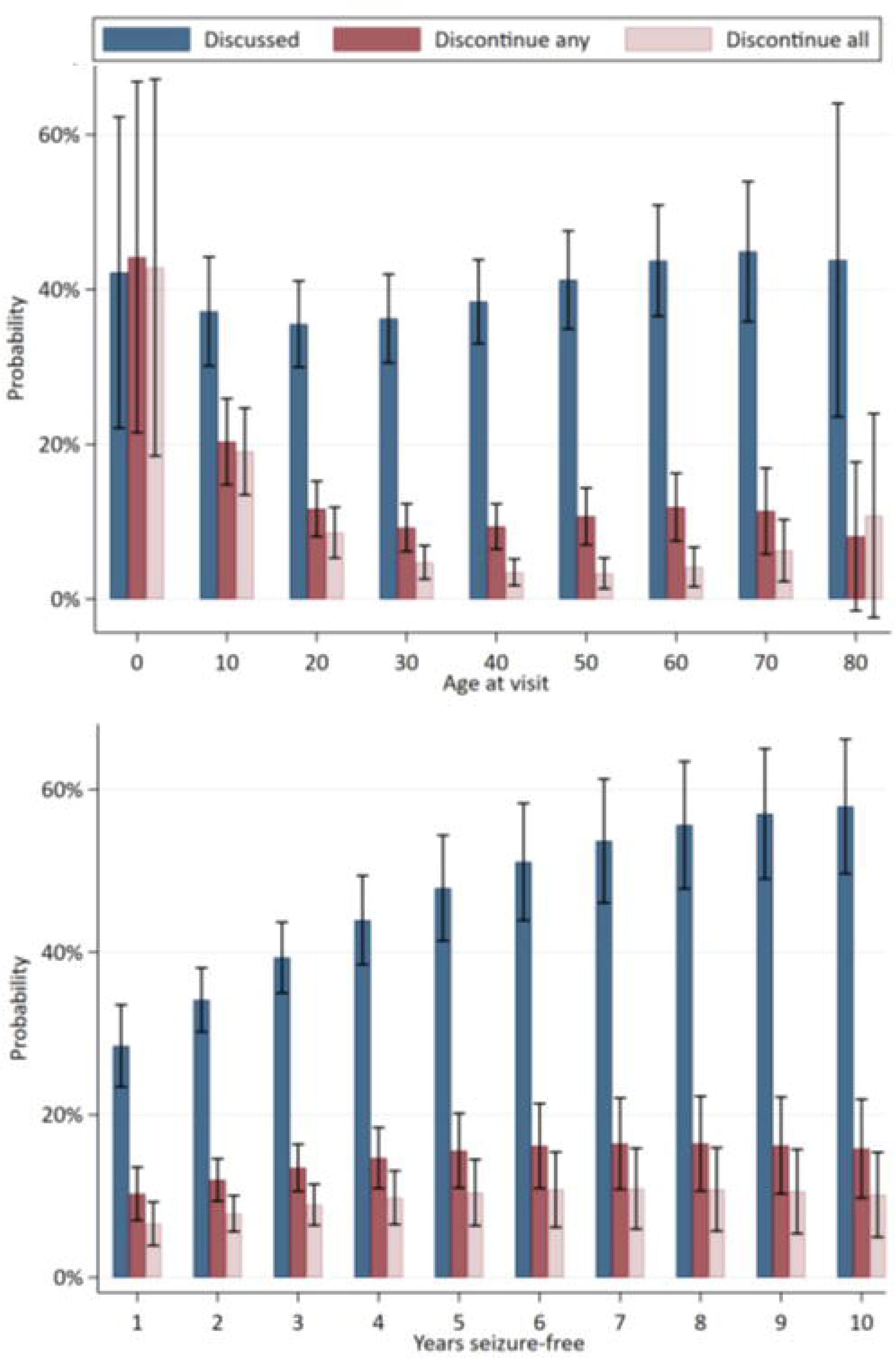
Unadjusted probabilities and 95% confidence intervals of discussing or planning to discontinue any or all antiseizure medications, stratified by a priori clinical variables. Further unadjusted percentages and p-values may be found in Table 2. Each visit represented an observation.

**Supplemental Figure 2:**
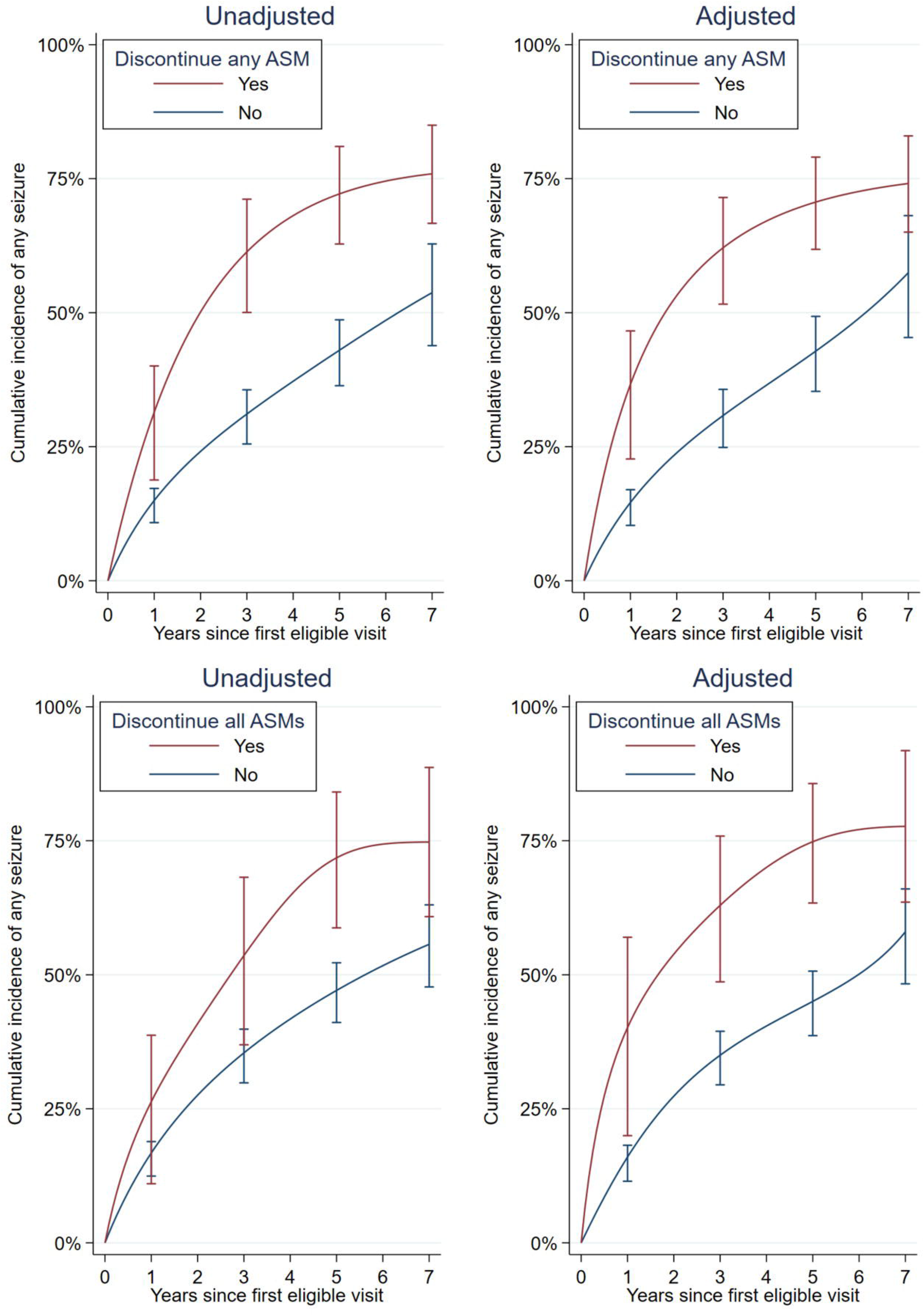
Time to first seizure relapse (95% confidence intervals) according to whether patients planned to discontinue at least one (top) or all (bottom) antiseizure medications (ASMs). The origin of time in this graph is the date of the first visit in our study period. Adjusted curves were adjusted for all variables listed in Table 2. Confidence intervals are spaced every two years below for display.

## Methods Supplement

We calculated seizure relapse curves as follows. First, we assessed the date of the patient’s first visit (beginning follow-up 1/2015), the date when each patient decided to discontinue if applicable, the date of the patient’s first seizure prior to January 1, 2022, if any, and the date of the patient’s last follow-up visit prior to January 1, 2022. We included one observation for each person-month of follow-up. The main predictor was whether the patient decided to discontinue ASMs, which started out as “no,” until the first instance of deciding to discontinue if applicable, after which this variable changed to “yes” for the remainder of follow-up. We chose ‘planned discontinuation’ as the main predictor rather than ‘completion of discontinuation,’ given ‘completion’ would have suffered from immortal time bias (only the lowest risk patients complete tapering without having a seizure). Patients were censored upon their first seizure after their first eligible visit, their last follow-up visit, or January 1, 2022, whichever came first. We then calculated an unadjusted parametric survival curve from a discrete time logistic model, then a standardized survival curve adjusted for all covariates listed in Table 2 in accordance with best practices.^39^ We calculated cumulative incidence as 1 minus the survival curve at each timepoint. We obtained confidence intervals via 1,000 bootstrapped replications. This approach provided several key advantages: 1) encoded tapering as a time-varying covariate (to avoid misattributing pre-discontinuation time to the “discontinuation” group), 2) avoided requiring the proportional hazards assumption which applies to Cox models but not discrete time logistics models, and 3) adjusted survival curves which otherwise would not have been possible if we had simply drawn Kaplan-Meier curves.

